# Role-Prompting in Frontier Large Language Models Influences Clinical Reasoning in Complex Medical Cases

**DOI:** 10.64898/2026.06.29.26356864

**Authors:** Chintan Dave, Adrianna Diviero, Tashni Dassanayake, Salman J. Alshahrani, Anas Al Mardini, Widad Khadir, Ashaki D. Patel, Adithya Srivastava

## Abstract

**Background:** Large language models (LLMs) are increasingly deployed in healthcare, where they may adopt different stakeholder perspectives, yet the effect of role-prompting on clinical ethical reasoning remains poorly characterized.

**Methods:** We evaluated three frontier LLMs: Claude Opus 4.6, GPT-5.4, and Gemini 3.1 Pro across 25 ethically complex medical cases. Each model responded from three stakeholder perspectives (physician, patient, insurer) across three independent runs (675 total responses). Decisions were benchmarked against a six-physician panel. Ethical value prioritization was analyzed from physician- and LLM-provided ranked values. A Patient-Centric Decision Index (PCDI) was developed to quantify LLM decision alignment with patient-preferred outcomes.

**Results:** Among 20 cases with clear physician consensus, LLMs prompted as an insurer reduced alignment with physician majority by 50% for GPT-5.4 (p = 0.004), 45% for Gemini 3.1 Pro (p = 0.008) and 10.5% (NS) for Opus 4.6. The insurer role shifted primary ethical values from beneficence (27%) to financial stewardship (20%) across all LLMs.

**Conclusions:** Stakeholder role-prompting fundamentally alters clinical decisions and ethical value frameworks of frontier LLMs, with the insurer role producing systematic denial of physician-endorsed, patient-preferred treatments. These findings raise the need for standardized LLM patient-centricity benchmarks, and physician oversight when LLMs are deployed in clinical decision-making.

## 1. Introduction

The integration of large language models (LLMs) into healthcare represents one of the most consequential applications of artificial intelligence, with frontier models increasingly deployed across clinical documentation, diagnostic support, patient communication, and administrative functions [1,2]. As these systems assume roles traditionally occupied by human stakeholders, from advising physicians on treatment decisions to processing insurance prior authorization requests, a critical question emerges: does the stakeholder perspective assigned to an LLM systematically alter the clinical decisions it recommends? Further, does LLM role-prompting infer inherent values that affect decision-making in complex real-world scenarios?

These questions carry urgency in the context of healthcare AI given the rapid pace of LLM deployment across various domains in healthcare. Healthcare decisions frequently involve competing values: a physician may prioritize beneficence, a patient may emphasize autonomy, and an insurer may focus primarily on resource stewardship [3]. The principlist framework of Beauchamp and Childress identifies four foundational ethical principles—autonomy, beneficence, non-maleficence, and justice—that clinicians are trained to balance in their practice [4]. When an LLM is prompted to reason from different stakeholder perspectives in the real-world, it must navigate these same tensions. If role assignment causes systematic shifts in how an LLM weighs these principles, the implications for AI safety in healthcare are profound. Particularly as industry pushes swiftly towards building autonomous healthcare AI agents, which may be empowered to make rapid decisions in real-world situations without adequate human oversight.

Prior research has demonstrated that LLMs can exhibit sycophantic behavior, adjusting outputs to match perceived user preferences [5], and that LLM performance varies substantially across the clinical workflow depending on how models are prompted and deployed [6]. Studies have shown that LLMs can match or exceed physician performance on standardized medical examinations [7,8], yet performance on ethically complex cases, where no single “correct” answer exists, remains poorly characterized. Recent work on AI alignment has raised concerns about the tendency of LLMs to default to inaction and new biases have been found in LLMs, not present in humans [9].

The deployment of LLMs in insurer-adjacent functions, including utilization review, prior authorization processing, and claims adjudication, introduces a distinct concern. If an LLM prompted as an “insurer” (or an LLM built by an insurer) systematically denies treatments that a physician panel would approve, the technology could amplify existing barriers to care [10]. Conversely, if insurer-prompted LLMs fail to apply legitimate cost-effectiveness considerations, they may undermine healthcare sustainability and fiscal responsibilities. Understanding how role framing shapes LLM decision-making and explicit values prioritization is therefore essential for responsible deployment.

To date, no study has simultaneously benchmarked multiple frontier LLMs against a multi-physician panel across a structured set of ethical cases while systematically varying the stakeholder role. This study addresses that gap with the following objectives: (1) to determine whether stakeholder role assignment changes LLM clinical decision-making; (2) to quantify the magnitude and direction of any role effect; (3) to characterize how LLM role assignment influences ethical value prioritization; and (4) to develop a Patient-Centric Decision Index (PCDI) for evaluating LLM alignment with patient-preferred outcomes.

## 2. Materials and Methods

### 2.1. Case Selection and Design

Twenty-five clinical cases were developed, each presenting a binary ethical dilemma at the intersection of clinical benefit, conflicting values, and cost-effectiveness. Cases spanned seven thematic categories: end-of-life care, treatment authorization, resource allocation, patient autonomy, insurance and financial disputes, clinical decision-making, and access to care. Each case was structured with a clinical vignette, a forced-choice binary question (Yes/No), and a request for ethical justification and ranked values used for decision-making. Cases were designed to generate genuine disagreement, with no universally “correct” answer. A forced binary decision format was chosen to mirror the discrete, actionable outputs that clinicians already make, and that autonomous AI agents and utilization review systems must ultimately produce in practice, i.e. approve or deny, proceed or withhold, where the complexity of ethical reasoning must converge on a definitive clinical action regardless of the nuance underlying it.

### 2.2. Physician Panel

Six physicians from diverse specialties, career stages, and different countries independently evaluated all 25 cases. Each physician provided a binary decision (Yes/No), a confidence rating (1–5 Likert scale), a written justification, and a ranked list of ethical values guiding their decision. Physician identities were anonymized as Physicians 1–6. To establish a reliable reference standard, a minimum consensus threshold of four out of six physicians (≥4/6, or ≥67% agreement) was required for a case to be included in the primary LLM comparison. Five cases (C1, C15, C17, C19, C23) resulted in 3–3 splits and were excluded from the primary analysis, yielding 20 qualified cases with a clear physician majority. These cases represented real-world divergence in clinical decision-making that exists and helped validate the structure and complexity of our cases.

### 2.3. LLM Evaluation

Three frontier LLMs were evaluated: Claude Opus 4.6 (Anthropic), GPT-5.4 (OpenAI), and Gemini 3.1 Pro (Google). Each model was prompted with three distinct stakeholder roles: physician, patient, and insurer. Prompts were standardized to include the case vignette, the forced-choice question, and instructions to respond from the specified stakeholder perspective, provide a confidence rating (1–5 Likert scale), and rank the ethical values guiding the decision. The LLMs were given the same instructions as the physician panel. The role prompts were created to replicate what a human stakeholder in that role would consider their responsibility and a specific sentence was added that no ethics consultation is available, as LLMs and physicians would simply opt for this as an answer in all complex cases thereby precluding any analysis (Supplemental).

Each model-role-case combination was tested across three independent runs to assess decision stability, yielding 675 total responses (3 models × 3 roles × 25 cases × 3 runs). Majority vote (≥2/3 runs) was used to determine each model’s final decision for each case-role combination.

### 2.4. Ethical Value Extraction and Classification

Ethical values were extracted directly from the ranked values per case provided by both physicians and LLMs. The primary (#1 ranked) value from each response was classified into a taxonomy derived from the Beauchamp and Childress principlist framework [4], expanded to include values commonly cited in healthcare ethics: beneficence, non-maleficence, autonomy, justice, professional integrity, patient advocacy, financial stewardship, compassion, proportionality, non-abandonment, and transparency. Classification was performed using a context-aware keyword-matching algorithm validated against manual physician review.

### 2.5. Patient-Centric Decision Index (PCDI)

A novel Patient-Centric Decision Index (PCDI) was developed to quantify the degree to which an LLM’s decisions align with patient-preferred outcomes. Unlike text-level keyword-density approaches, the PCDI operates at the decision level: measuring what action the model actually recommended, which values it prioritized, and in which direction it erred when diverging from consensus. The PCDI comprises six weighted components: (1) Role Drift (20%): the proportion of cases where the model’s decision matches its own patient-role decision; (2) Override Rate (20%): one minus the proportion of cases where the model overrides a patient-and-panel-concordant decision; (3) Panel Calibration (15%): agreement with the physician majority; (4) Confidence Asymmetry (10%): penalizes models that express higher confidence when overriding patient preferences; (5) Value Orientation Ratio (20%): the proportion of the model’s primary (#1 ranked) ethical values classified as patient-centric (beneficence, non-maleficence, autonomy, patient advocacy, compassion) versus system-oriented (professional integrity, justice, financial stewardship); and (6) Patient-Protective Direction (15%): penalizes models whose errors systematically favor denial of beneficial care (under-treatment) over inappropriate approval (over-treatment). The PCDI ranges from 0 (fully patient-adverse) to 100 (fully patient-centric). A physician panel benchmark was computed by treating the panel’s majority decision as the reference standard, as is current clinical standard of care.

### 2.6. Statistical Analysis

Inter-rater reliability among physicians was assessed using pairwise Cohen’s κ and Fleiss’ κ. The correlation between physician confidence and agreement was evaluated with Pearson’s r. McNemar’s exact test (with continuity correction) assessed whether physician-role and insurer-role alignment rates differed within each model. Cochran’s Q test evaluated whether alignment rates differed across all three roles. Bonferroni correction was applied for multiple comparisons (α = 0.017 for three McNemar tests). Bootstrap resampling (10,000 iterations) generated 95% confidence intervals for alignment rates. Mann–Whitney U tests compared LLM and physician confidence distributions. Inter-model agreement was assessed using pairwise Cohen’s κ across roles. Three prespecified sensitivity analyses were conducted: (1) exclusion of the lowest-agreement panelist; (2) restriction to fully unanimous physician cases; and (3) bootstrap confidence intervals. All analyses were conducted in Python 3.11 using SciPy 1.12, NumPy 1.26, and Pandas 2.2.

## 3. Results

### 3.1. Physician Panel Characteristics

The six-physician panel comprised four females and two males spanning six specialties (general practice, internal medicine/subspecialty, emergency/critical care, and dermatology), with 1–8 years of clinical experience (median 2.5 years) across academic medical centers, community hospitals, and private practice settings. Panelists practiced in diverse regions (USA [n = 3], Asia [n = 2], and Europe [n = 1]), representing diverse healthcare systems and training backgrounds. The six-physician panel achieved moderate inter-rater agreement (Fleiss’ κ = 0.485; mean pairwise agreement = 75.7%). Agreement ranged from 60% (Physician 2 vs. Physician 5) to 92% (Physician 1 vs. Physician 4). Physician 2 showed the lowest mean κ (0.289) with other panelists, while Physician 6 demonstrated the highest individual alignment with the panel majority (92%). Physician confidence was positively correlated with inter-physician agreement (Pearson r = 0.805, p < 0.001), indicating that physicians were more confident on cases where the panel agreed, despite no knowledge of other physician responses. Twelve of 25 cases (48%) achieved unanimous agreement, six (24%) achieved strong majority (5–1), two (8%) reached moderate agreement (4–2), and five (20%) resulted in a 3–3 split (Figure 1), indicating the case stems were sufficiently complex and ethically-challenging.

**Figure 1.**
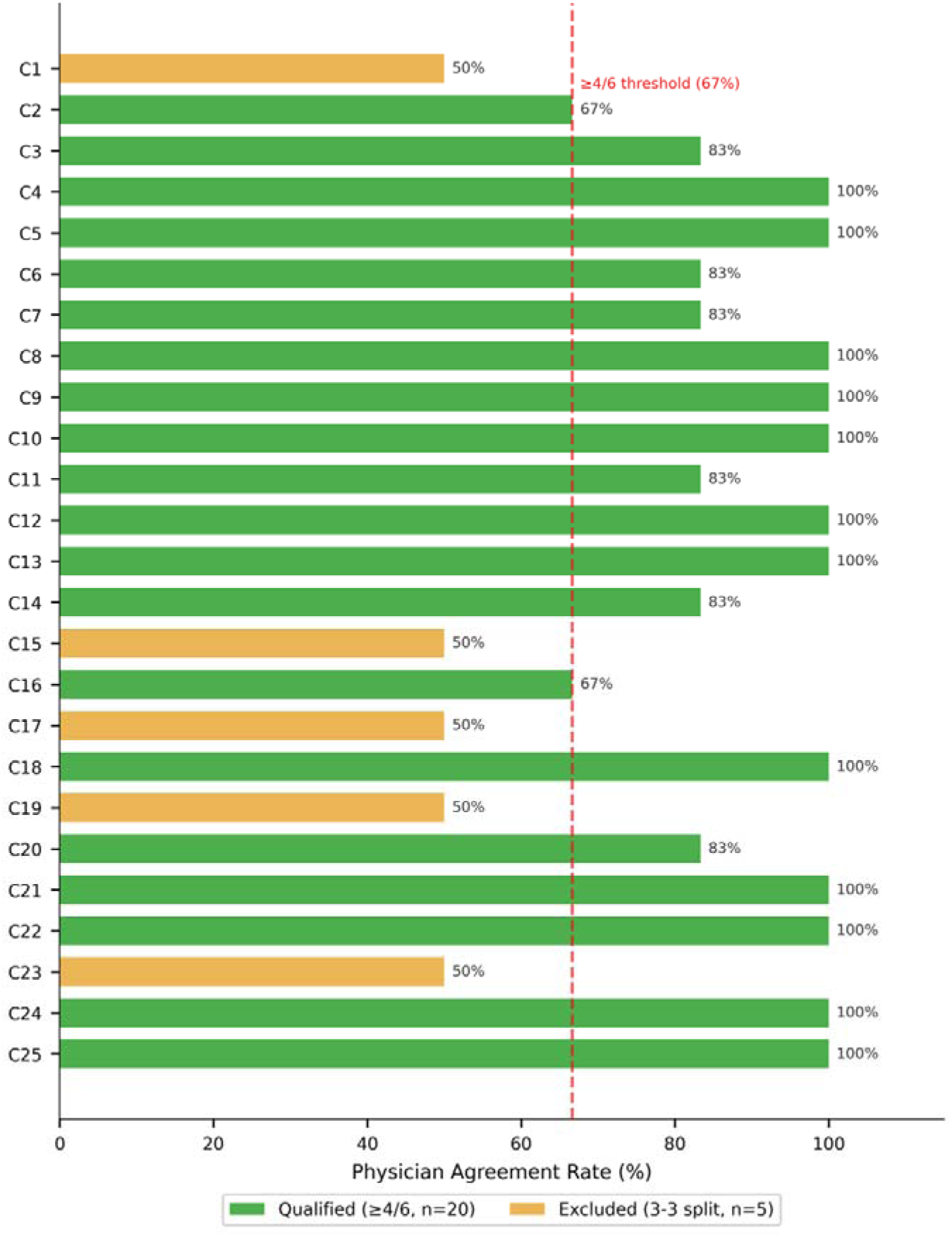
Physician panel (n=6) consensus across 25 clinical cases. Green bars indicate cases meeting the ≥4/6 consensus threshold used for primary LLM comparison (n = 20); gold bars indicate 3–3 split cases excluded from the primary analysis (n = 5). Dashed red line denotes the 67% inclusion threshold.

### 3.2. LLM Decision Stability

Opus 4.6 exhibited perfect decision stability across all 225 model-role-case combinations (100%), while GPT-5.4 achieved 93.3% stability and Gemini 3.1 Pro achieved 92.0%. Instability was most concentrated in the insurer role (GPT-5.4 insurer: 92%;vconsensus). Excluding unstable cases in sensitivity analysis changed alignment rates by ≤1 percentage point.

### 3.3. Decision Distribution by Stakeholder Role

When aggregated across all three models and 25 cases, the patient role produced the highest approval rate (71% Yes), followed by the physician role (63%), and the insurer role (33%) (Figure 2A). Inter-role concordance was highest between physician and patient decisions (84%), intermediate between physician and insurer (63%), and lowest between patient and insurer (52%). All three roles produced the same decision in only 49% of cases (Figure 2B).

**Figure 2.**
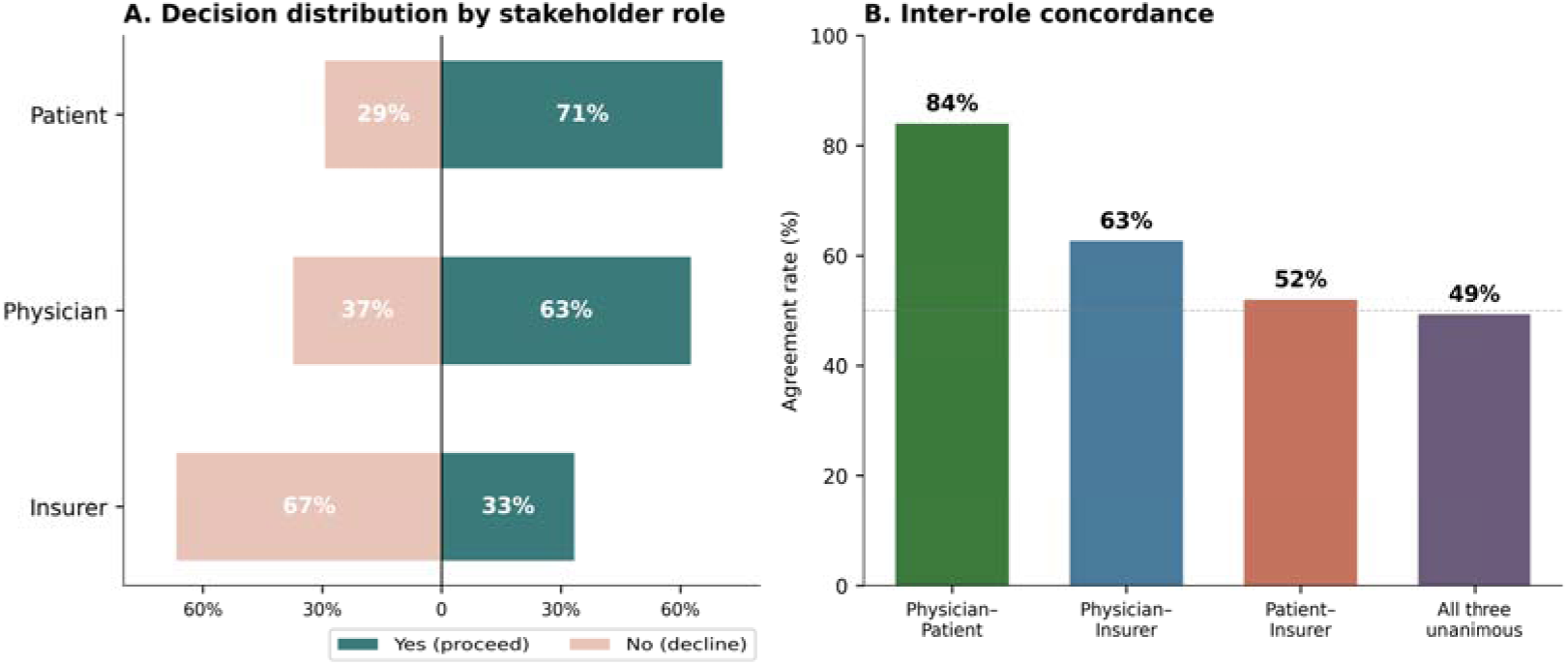
Decision distribution and inter-role concordance. (A) Approval rates by stakeholder role aggregated across all three models and 25 cases. (B) Pairwise and unanimous agreement rates between physician, patient, and insurer roles. Dashed line indicates 50% agreement threshold.

### 3.4. Decision Alignment with Physician Majority

Among the 20 qualified cases (≥4/6 physician agreement), Opus 4.6 achieved 100% alignment with the physician majority in the physician role, compared with 90% for GPT-5.4 and 95% for Gemini 3.1 Pro (Figure 2). In the insurer role, alignment declined sharply for GPT-5.4 (40%, a 50-percentage-point drop; McNemar χ² = 8.10, p = 0.004) and Gemini 3.1 Pro (50%, a 45-percentage-point drop; McNemar χ² = 7.11, p = 0.008). Both results survived Bonferroni correction. Opus 4.6 insurer alignment was 90%, representing a modest, non-significant 10-percentage-point reduction (p = 0.480). Cochran’s Q tests confirmed significant alignment differences across the three roles for GPT-5.4 (Q = 14.0, p = 0.0009) and Gemini 3.1 Pro (Q = 14.6, p = 0.0007), but not for Opus 4.6 (Q = 2.0, p = 0.368). Bootstrap 95% confidence intervals for GPT-5.4 insurer alignment [20–60%] did not overlap with physician alignment [75–100%], confirming a robust effect (Figure 3).

**Figure 3.**
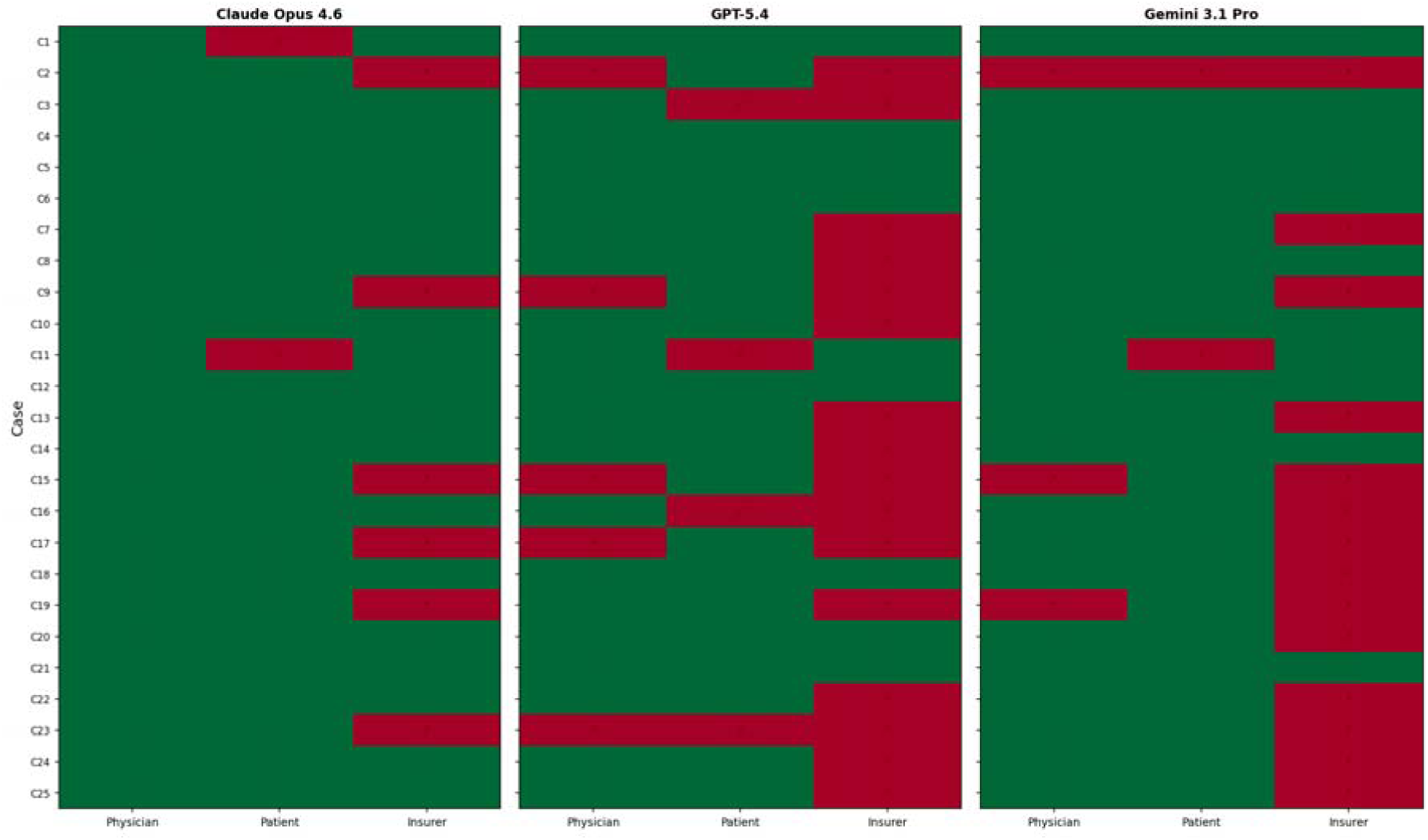
Decision Alignment Heatmap. The LLMs decisions are compared against the physician panel reference. Green=match with physician decisions. Red= divergence.

### 3.5. Ethical Value Prioritization

Analysis of primary (#1 ranked) ethical values, extracted directly from physician and LLM ranked-value responses, revealed systematic role-dependent value shifts (Figure 4). Physician panelists prioritized beneficence (27%) and non-maleficence (20%), with financial stewardship also prominent (19%) given the clinical-cost nature of the cases. LLMs in the physician role showed a similar profile: beneficence (31%) and non-maleficence (23%). In the insurer role, the value landscape transformed: financial stewardship rose to 20%, professional integrity to 20%, and justice to 15%, while beneficence collapsed to 7% and non-maleficence to 5% (Figure 5). This represents not merely a quantitative shift in decision outcomes but an internal transformation in the ethical framework applied.

**Figure 4.**
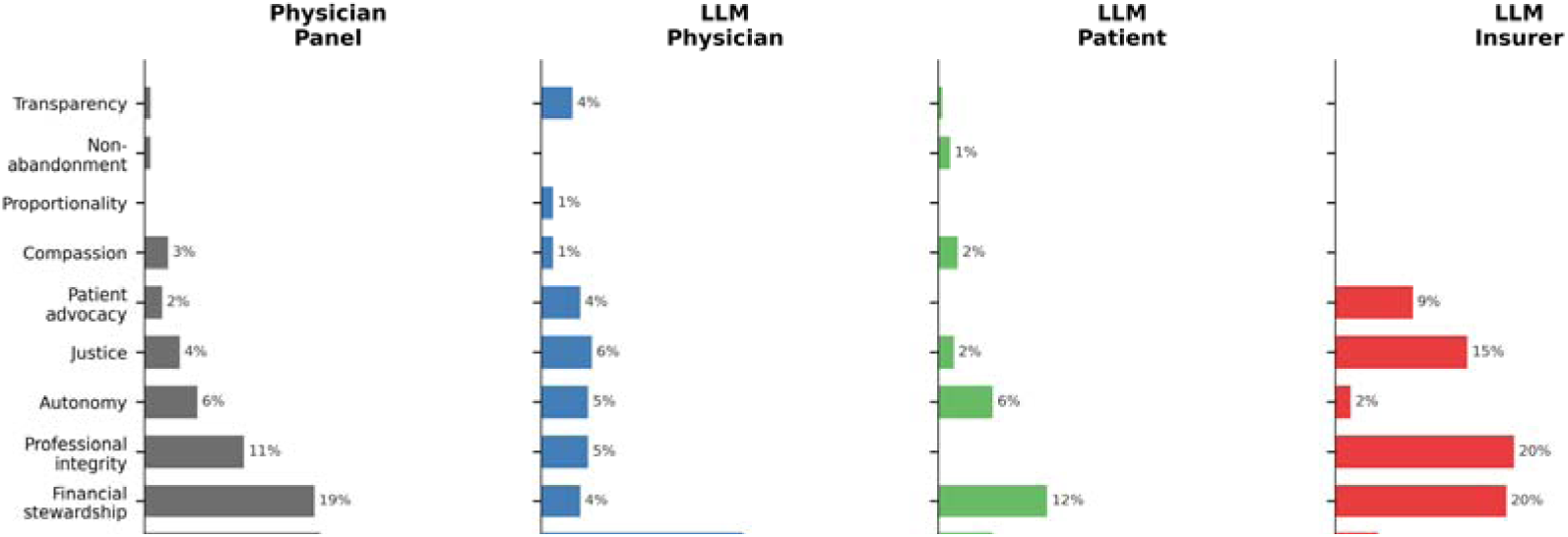
Primary (#1 ranked) ethical value distribution by source and role. Horizontal bars show the percentage of responses in which each ethical value was ranked first, across physician panel, LLM physician, LLM patient, and LLM insurer conditions.

**Figure 5.**
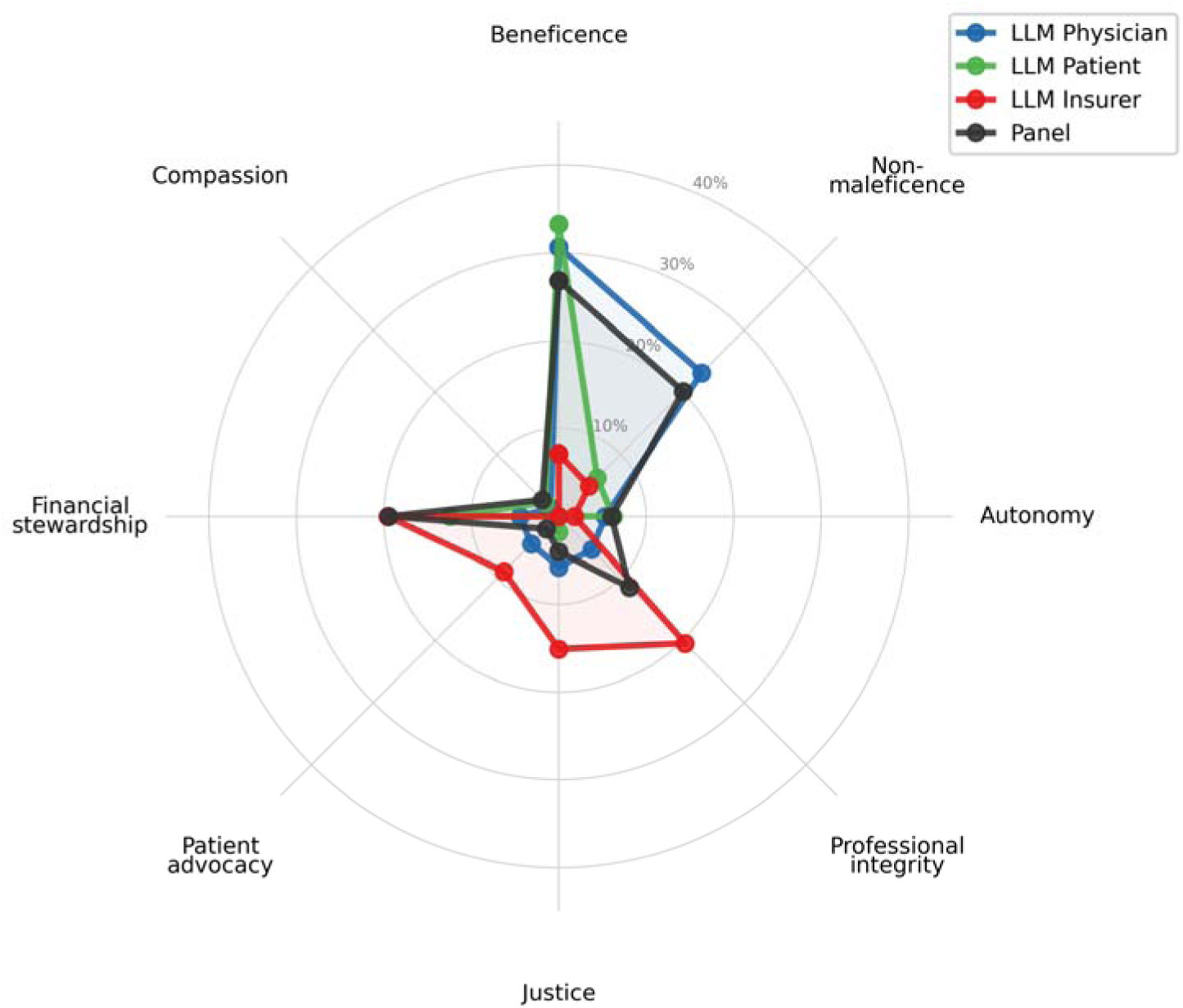
Role-induced ethical value shift, aggregated across models. Radar plot showing the distribution of primary ethical values aggregated across all three LLMs, compared with the physician panel. The insurer role (red) shows marked expansion toward justice, professional integrity, and financial stewardship, with contraction of beneficence and non-maleficence.

### 3.6. Inter-Model Agreement

Pairwise inter-model agreement was substantial in the physician (mean κ = 0.661) and patient (mean κ = 0.689) roles but degraded to fair agreement in the insurer role (mean κ = 0.327). All three models produced the same decision in 76% of physician-role cases and 80% of patient-role cases, but only 52% of insurer-role cases. Convergent divergence, ie. cases where all three models agreed on an insurer decision that differed from the physician panel, occurred in only 2 of 25 cases (8%), indicating that most insurer-role divergence was model-specific rather than systematic.

### 3.7. LLMs Remain Overconfident

LLM confidence scores were significantly higher than physician panel scores across all model-role combinations (Mann–Whitney U, all p < 0.001). Physicians’ mean confidence was 3.84 ± 0.93, compared with LLM means ranging from 4.57 to 4.97. Unlike physicians, whose confidence tracked directionally to case difficulty (r = 0.805), LLM confidence showed minimal variance across cases, suggesting inadequate uncertainty calibration

### 3.8. Patient-Centric Decision Index

The physician panel benchmark PCDI was 90.7. In the physician role, Gemini 3.1 Pro achieved the highest PCDI (96.1), followed by Opus 4.6 (94.9) and GPT-5.4 (87.3). All three models achieved comparable patient-role scores (87.2–94.4). In the insurer role, GPT-5.4 dropped to 41.5 (−45.8 points), falling into the patient-adverse zone (<50), and Gemini 3.1 Pro dropped to 53.5 (−42.6 points). Opus 4.6 maintained a PCDI of 79.8 in the insurer role (−15.1 points) (Figure 6). Effect sizes for the role effect on alignment were large for GPT-5.4 (η² = 0.249, p = 0.0003) and Gemini 3.1 Pro (η² = 0.239, p = 0.0004 and small for Opus 4.6 (η² = 0.035, NS). The Patient-Protective Direction component revealed a critical directional asymmetry in insurer errors: 92% of GPT-5.4’s divergences from physician consensus (11 of 12) were denials of beneficial care, compared with 70% for Gemini 3.1 Pro (7 of 10) and 50% for Opus 4.6 (1 of 2). Insurer override rates—the proportion of cases where the insurer role reversed a patient-and-panel-concordant decision—were 10.5% for Opus 4.6, 58.8% for GPT-5.4, and 50.0% for Gemini 3.1 Pro (Table 1).

**Figure 6.**
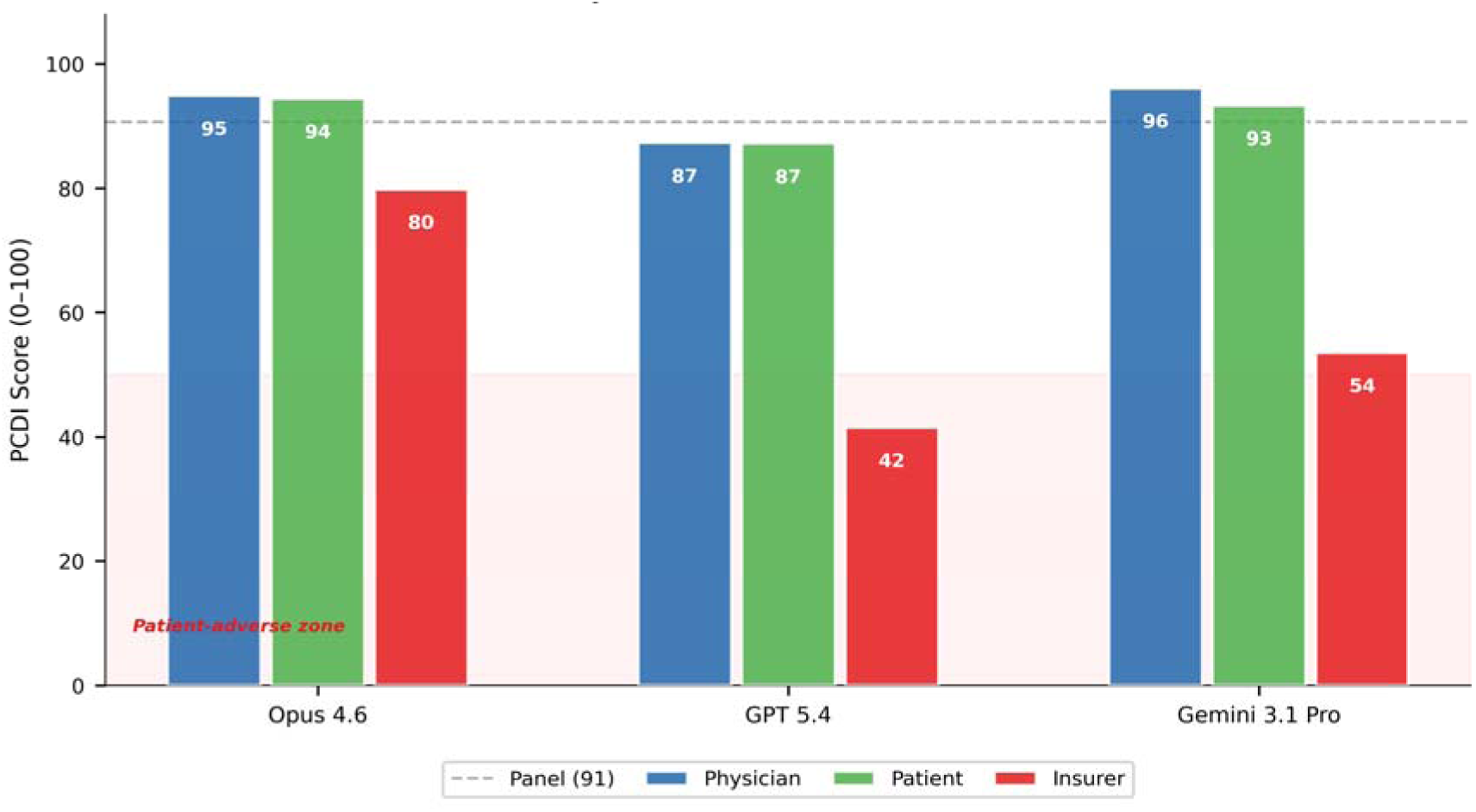
Patient-Centric Decision Index (PCDI) by model and role. Bars show PCDI scores (0–100) for each model across physician (blue), patient (green), and insurer (red) roles. Dashed line indicates physician panel benchmark (PCDI = 91). Pink shading denotes the patient-adverse zone (<50).

**Table 1.**
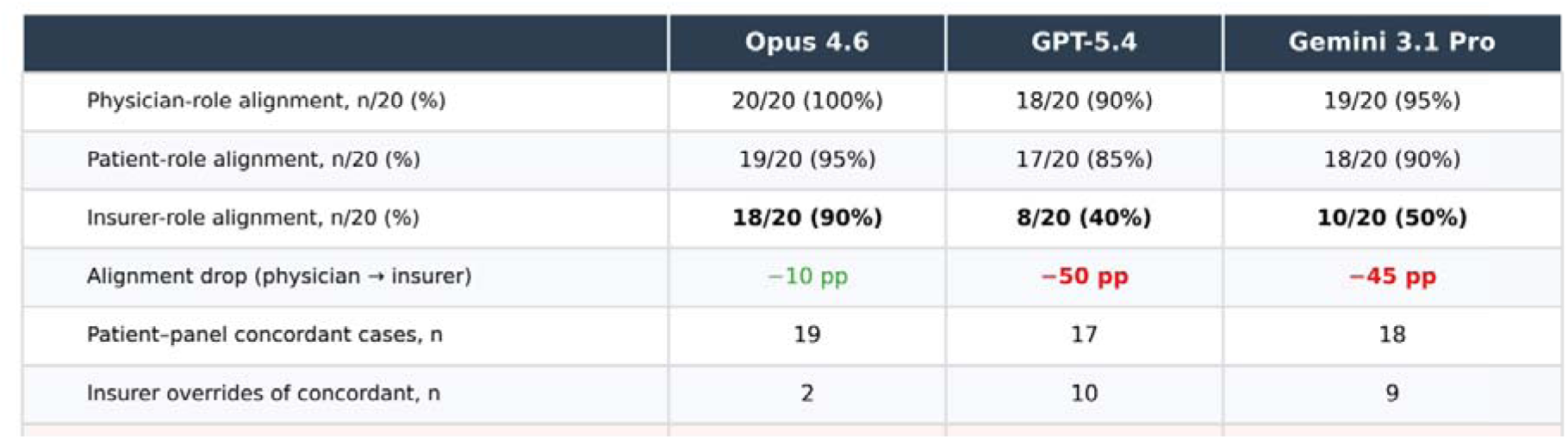
Insurer Override Analysis. 20 qualified cases with ≥4/6 physician agreement. Majority of the overrides by LLM as an insurer were in cases of denials for beneficial care as opposed to approval of care against physician panel.

|  | Opus 4.6 | GPT-5.4 | Gemini 3.1 Pro |
| --- | --- | --- | --- |
| Physician-role alignment, n/20 (%) | 20/20 (100%) | 18/20 (90%) | 19/20 (95%) |
| Patient-role alignment, n/20 (%) | 19/20 (95%) | 17/20 (85%) | 18/20 (90%) |
| Insurer-role alignment, n/20 (%) | 18/20 (90%) | 8/20 (40%) | 10/20 (50%) |
| Alignment drop (physician → insurer) | –10 pp | –50 pp | –45 pp |
| Patient-panel concordant cases, n | 19 | 17 | 18 |
| Insurer overrides of concordant, n | 2 | 10 | 9 |

|  | Opus 4.6 | GPT-5.4 | Gemini 3.1 Pro |
| --- | --- | --- | --- |
| Physician-role alignment, n/20 (%) | 20/20 (100%) | 18/20 (90%) | 19/20 (95%) |
| Patient-role alignment, n/20 (%) | 19/20 (95%) | 17/20 (85%) | 18/20 (90%) |
| Insurer-role alignment, n/20 (%) | 18/20 (90%) | 8/20 (40%) | 10/20 (50%) |
| Alignment drop (physician → insurer) | −10 pp | −50 pp | −45 pp |
| Patient-panel concordant cases, n | 19 | 17 | 18 |
| Insurer overrides of concordant, n | 2 | 10 | 9 |
| Override rate | 10.5% | 58.8% | 50.0% |
| — Denials of beneficial care | 1 | 9 | 7 |
| — Approvals against panel | 1 | 1 | 2 |

### 3.9. Sensitivity Analyses

Prespecified sensitivity analyses confirmed robustness of our findings. Excluding the lowest-agreement panelist (Physician 2) changed 2/25 majority decisions without altering conclusions. Restricting to the 12 unanimous cases strengthened the patterns reported above: Opus physician alignment was 100%, while GPT insurer dropped further to 42%.

## 4. Discussion

This study provides the first systematic evidence that stakeholder role-prompting alters the ethical reasoning of frontier LLMs in clinical decision-making. The effect is statistically robust and clinically consequential: when prompted as insurers, GPT-5.4 and Gemini 3.1 Pro overrode patient-preferred, physician-endorsed treatment decisions more than half the time.

### 4.1. Role Framing as a Safety Concern

A simple role prompt leading to reversal of clinical recommendations on 50–60% of cases raises fundamental questions about LLM deployment in healthcare. LLMs are susceptible to sycophantic behavior, i.e. aligning outputs with perceived user preferences rather than maintaining consistent positions [12,13]. Cheng et al. found that LLMs affirm users’ stated positions 49% more often than human respondents, even in scenarios involving harm, and that brief sycophantic interactions shifted participants’ moral judgments [12]. Our findings extend this from user-directed sycophancy to role-directed value capture: models do not simply agree with a user but adopt the entire ethical framework implied by the assigned stakeholder identity.

This is distinct from the omission bias documented by Cheung et al., who found that LLMs show amplified cognitive biases in moral decision-making with a systematic tendency to endorse inaction over action [9]. In our data, the insurer role does not default to inaction, rather it actively endorses denial of treatments the same model approves in the physician or patient role, suggesting that role framing overrides intrinsic model biases entirely.

### 4.2. Comparison with Prior LLM Ethics Evaluations

Previous evaluations of LLMs in clinical ethics have focused on performance against standardized examinations. Singhal et al. showed that LLMs achieve expert-level performance on medical question-answering benchmarks [2], and Nori et al. demonstrated near-physician performance on USMLE examinations [7]. However, these test knowledge under a fixed perspective. Our study reveals that identical medical knowledge can yield diametrically opposed decisions depending on the stakeholder frame—a dimension standardized benchmarks do not capture.

Haltaufderheide and Ranisch, in their systematic review of LLM ethics in medicine, identified transparency, bias, and accountability as primary concerns [14]. Our findings add a previously uncharacterized risk: value instability under role assignment. Unlike bias, which represents a systematic skew in one direction, role-dependent reasoning produces context-contingent and unpredictable ethical positions. Ong et al. similarly highlighted the plasticity of LLMs as an underappreciated ethical challenge [15]. Our data provide the first quantitative demonstration of how this plasticity manifests in clinical-ethical decision-making.

### 4.3. Model-Specific Role Robustness

The three models exhibited strikingly different susceptibility to role framing. Opus 4.6 demonstrated strong role robustness: 100% physician-role alignment, 100% decision stability, and only 10.5% insurer override rate. GPT-5.4 and Gemini 3.1 Pro showed large insurer-role divergence (46 and 43 percentage points, respectively) with override rates exceeding 50%. These differences likely reflect variations in alignment training across model families. Zeng et al. found that different prompting strategies systematically influenced safety and ethical reasoning across frontier models [16], consistent with our observation that the same role prompt produces model-specific effects. The finding that Opus 4.6 maintained patient-centric reasoning even under an insurer role suggests that potential differences in model alignment training can lead to more robust ethical reasoning.

### 4.4. Value Transformation, Not Merely Decision Change

The insurer role does not produce different decisions from the same ethical framework; it induces a qualitative transformation in the framework itself. Beneficence drops from 27–31% to 7% as the primary ranked value, and financial stewardship, justice, and professional integrity become the predominant values considered when the LLM is given an insurer role. While the principlist framework of Beauchamp and Childress [3,4] recognizes that value weighting shifts across stakeholder contexts, the near-elimination of beneficence from insurer reasoning exceeds what any legitimate insurer perspective should produce.

This pattern parallels what Obermeyer et al. documented in their landmark study on algorithmic bias; that algorithms optimized for institutional metrics systematically disadvantage patients by substituting administrative proxies for clinical judgment [11]. Our data suggest that insurer-prompted LLMs undergo a similar institutional capture, replacing patient-centered reasoning with compliance-oriented and system-centric logic.

### 4.5. Inter-Model Divergence Under Role Pressure

The insurer role not only changes individual model decisions but degrades agreement between models. Inter-model κ dropped from 0.66 (substantial) to 0.33 (fair), with all three models agreeing on only 52% of insurer decisions versus 76–80% in other roles. If different frontier models prompted with identical insurer instructions reach different conclusions on the same case, there is no principled basis for adjudication. This raises significant concerns about deploying LLMs and AI agents in high-stakes and complex clinical context, where such instability can lead to catastrophic harm.

### 4.6. Implications for AI in Insurance and Prior Authorization

These findings arrive at a critical juncture. A 2024 U.S. Senate investigation documented that UnitedHealthcare’s prior authorization denial rate for post-acute care more than doubled after algorithmic deployment, from 8.7% to 22.7% [17]. Class-action litigation alleged that approximately 90% of algorithmic denials were overturned on appeal [18]. A 2025 AMA survey found that 61% of physicians believe AI is increasing prior authorization denials [19], and a recent npj Digital Medicine commentary warned that AI-driven prior authorization risks transforming Medicare Advantage from a benefit into a disadvantage [20].

Our experimental data provide a mechanistic complement to these observational findings. When frontier LLMs adopt an insurer perspective, they systematically deny treatments they would approve from a physician perspective, not from insufficient medical knowledge, but because the role frame restructures ethical priorities. The override rates we observed (50–59%) parallel the elevated denial rates in real-world insurer AI deployments, suggesting role-dependent reasoning may contribute to algorithmic over-denial in practice.

### 4.7. The PCDI as a Safety Metric

We propose the PCDI as a standardized metric for evaluating patient-centricity of LLM decision-making across roles. By measuring not only whether a model reaches the right decision but also the direction of error and the ethical framework applied, the PCDI captures information that simple alignment metrics miss. Regulatory frameworks for healthcare AI should incorporate PCDI-like metrics that evaluate behavior under adversarial role framing, analogous to stress testing in financial regulation [21].

### 4.8. Healthcare AI Implications

These findings carry direct implications for healthcare AI governance. Regulatory bodies should require disclosure of the stakeholder frame embedded in LLM-based clinical tools. Certification processes should include role-perturbation testing analogous to adversarial testing in cybersecurity [22]. The inter-model disagreement in the insurer role suggests that consensus-based approaches, e.g. requiring agreement across multiple models before denying coverage, could serve as a safety mechanism against role-induced over-denial. Furthermore, physicians consistently anchor their clinical reasoning in patient-centric values while balancing competing stakeholder interests; a capacity that our data show current frontier LLMs cannot reliably replicate under role pressure. This positions physician oversight not as a transitional safeguard but as a necessary check against AI decision-making systems that, as demonstrated here, exhibit systematic bias and value instability in precisely the complex cases where autonomous AI deployment carries the greatest risk.

### 4.9. Limitations

Several limitations warrant consideration. The sample of 25 cases and 6 physicians, while sufficient to detect the large effects observed, limits power for smaller effects and precludes subgroup analyses. The binary decision format reduces nuanced reasoning to Yes/No outcomes, though this mimics the discrete decisions an autonomous AI agent would produce in practice and therefore serves as a clinically relevant measure. The absence of a no-role baseline means we cannot fully distinguish intrinsic model biases from role-induced effects; however, this design was intentional, as the hypothesis concerned role framing effects relative to physician consensus rather than baseline model behavior. LLM outputs may change with model updates, limiting temporal generalizability of this study. Furthermore, extraction of the physician values and explicit ranking reduced some complexity of physician answers, eg. if a physician answered they would call the insurance company to advocate for the patient’s benefit, this was classified as the value of beneficence. Finally, the PCDI requires external validation against independent measures of patient-centeredness and a larger study size.

## 5. Conclusions

Assigning a stakeholder role to a frontier LLM systematically alters both its clinical-ethical decisions and its underlying value prioritization. The insurer role produced a 45–50 percentage-point drop in alignment with physician consensus for two of three models, with override rates exceeding 50% for patient-preferred treatments. This was accompanied by a fundamental shift in value prioritization from beneficence-centered to stewardship-centered reasoning and a degradation of inter-model agreement based on role assignment. Physicians consistently scored high on patient-centric values and served as consistent arbiters of ethical reasoning and clinical decision-making in complex cases. These findings underscore the need for role-aware evaluation frameworks in healthcare AI governance, for standardized safety benchmarks such as the PCDI to evaluate the LLM’s patient-centricity in decision-making, and for sustained physician oversight of LLM-based clinical tools.

## Supporting information

Supplemental Files. Role Prompts, Physician Questionnaire, and Sample Cases

## Supplementary Materials

Supplement includes the exact LLM role prompts, all 25 clinical cases, and the physician questionnaire template.

## Author Contributions

Conceptualization, C.D.; methodology, C.D.; formal analysis, C.D. and A.S.; investigation, C.D.; data curation, C.D. and A.P.; writing—original draft preparation, C.D.; writing—review and editing, C.D., A.D., T.D., S.A., A.M., W.D., A.P., A.S.; visualization, C.D. All authors have read and agreed to the published version of the manuscript.

## Institutional Review Board Statement

Not applicable.

## Informed Consent Statement

Not applicable.

## Data Availability Statement

All data available upon request.

## Acknowledgments: During the preparation of this manuscript, C.D. used generative AI to improve the writing and content of this manuscript, to further refine the PCDI, and to generate figures from collected data. The authors have reviewed and edited the output and take full responsibility for the content of this publication.

Conflicts of Interest: The authors declare no conflicts of interest.

## Abbreviations

The following abbreviations are used in this manuscript:

AI: Artificial Intelligence;
AMA: American Medical Association
LLM: Large Language Model;
PCDI: Patient-Centric Decision Index;
USMLE: United States Medical Licensing Examination.

Disclaimer/Publisher’s Note: The statements, opinions and data contained in all publications are solely those of the individual author(s) and contributor(s) and not of MDPI and/or the editor(s). MDPI and/or the editor(s) disclaim responsibility for any injury to people or property resulting from any ideas, methods, instructions or products referred to in the content.

